# Prostatic Peripheral Zone Thickness: What is Normal on Magnetic Resonance Imaging?

**DOI:** 10.1101/2020.03.21.20039149

**Authors:** Neil F. Wasserman, Benjamin Spilseth, Tina Sanghvi

**Affiliations:** Department of Radiology, University of Minnesota Medical School

**Author notes:** Corresponding author Department of Radiology, Univ. of Minnesota Medical School, Mayo Mail Code 292, 420 Delaware Street S.E., Minneapolis MN 55455.

**Keywords:** Prostate, Volume, Peripheral Zone, Measurement, Methods

## Abstract

**Purpose:** To report the precision of a technique of measuring the PZ thickness on T2-weighted MRI and report normal parameters in patients with normal-sized prostates. We also wanted to establish the mean and second standard deviations (2SD) above and below the mean as criteria for abnormally narrow or expanded PZ thickness.

**Methods:** Of the initial 1566 consecutive cohort referred for evaluation for carcinoma based on elevated PSA (prostate specific antibody) or DRE (digital rectal examination), 132 separate subjects with normal-sized prostates were selected for this study. Mean age was 58.2 years (15-82). Median serum PSA was 6.2ng/ML (range, 0.3-145). Most were asymptomatic for lower urinary tract symptoms (LUTS). Inclusion criteria in this study required technically adequate T2-weighted MRI and total prostatic volume (TPV) ≤ 25 cc. Exclusion criteria included post prostatic surgical and radiation patients, patients having had medical management or minimally invasive therapy for BPH, those being treated for prostatitis. Patients with suspected tumor expanding or obscuring measurement boundaries were also not considered. Transition zone (TZ) and peripheral zone (PZ) volumes were determined using the prolate ellipsoid model. Postero-lateral measurement of the PZ was obtained at the axial level of maximal transverse diameter of the prostate on a line drawn from the outer boundary of the TZ to the inner boundary of the external prostatic capsule. The data was normally distributed. Therefore, it was analyzed using the 2-sided student t-test and Pearson produce correlation statistic.

**Results:** Mean pooled (composite) measurement for the postero-lateral PZ (PLPZ) was 10 mm (CI= 9.5-10.5 mm) with SD of 2.87 mm. Means were statistically the same for the 2 observers (p=0.75). Pearson correlation between the two observers was 0.63.

**Conclusions:** In a prostate ≤ 25 cc volume the postero-lateral PZ should be no thicker than 15.8 mm and averages 10.0 mm. when measured in the maximal axial plane on MRI. These norms were independent of age or use of endorectal coil. The technique measurement demonstrated clinically useful precision.

## Introduction

Prostatic issues are very common among males especially as they age. A lobar classification of benign prostatic hyperplasia (BPH) has been recommended for research studies evaluating strategies for treatment of BPH [1,2]. However, this classification does not include circumstances wherein the prostate is enlarged by digital rectal examination (DRE) or image-based total prostatic volume (TPV) measurement in the absence of significant imaged transition zone (TZ) nodular hyperplasia. This conundrum is exemplified by the following case:

> *A 71-year-old presented with mild obstructive voiding symptoms, a smooth non-tender mildly enlarged prostate on digital rectal examination (DRE) and prostate specific antigen (PSA) of 4*.*6 ng/mL. The clinical suspected diagnosis was” Probable benign prostatic hyperplasia (BPH). Rule out malignancy”*. *His T2-weighted MRI showed a TPV measuring 35*.*4 cc. suggesting enlargement and low signal areas in the peripheral zone (PZ). MRI interpretation was “BPH with suspicion of peripheral zone adenocarcinoma” (Fig. 1). Transrectal ultrasound-guided biopsies were negative for cancer*.

**Fig. 1.**
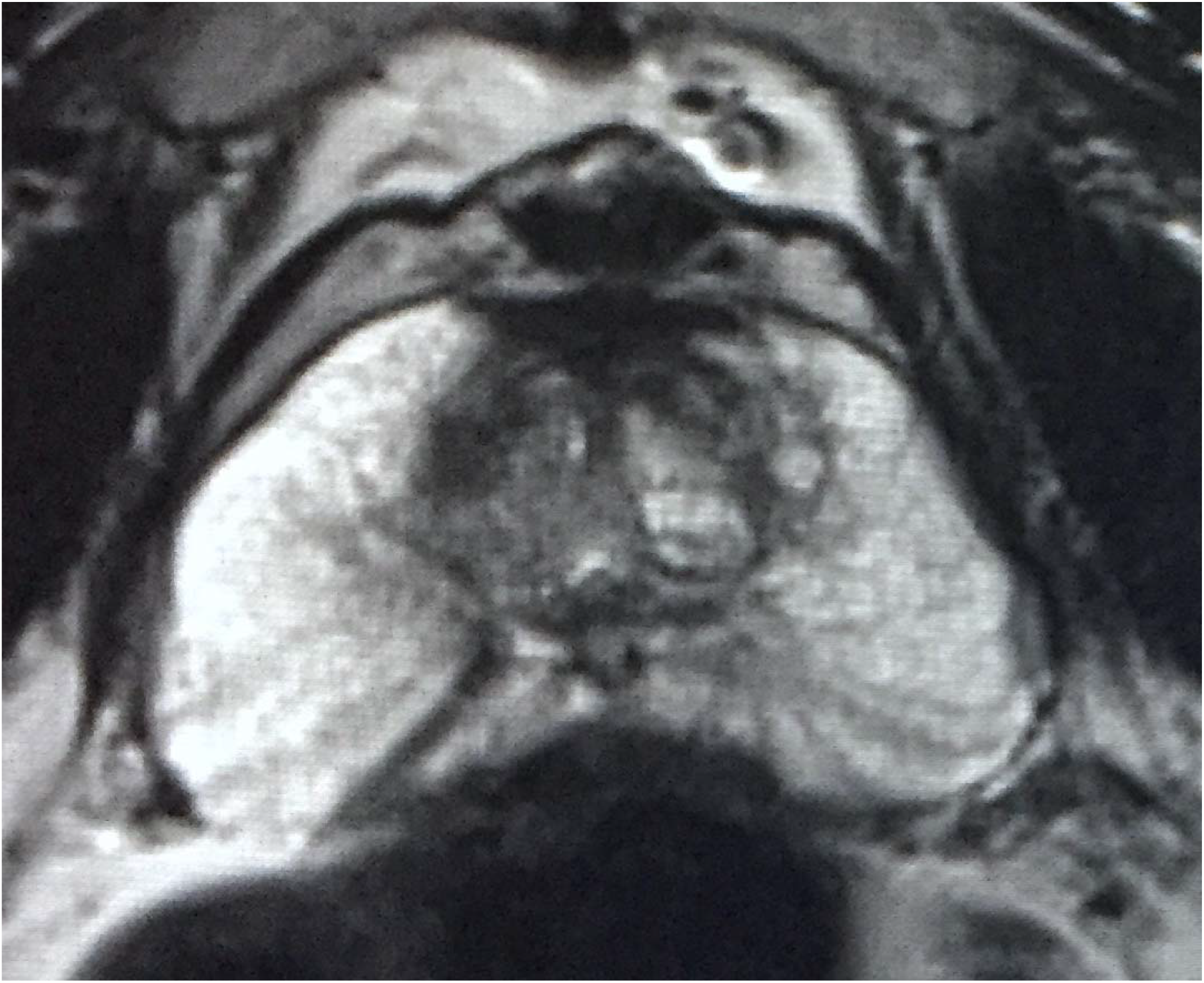
T2-weighted MRI on 71-year-old with “mild enlargement” on DRE and PSA of 4.6 ng/ml. Total prostatatic volume measured 35.4 cc. TZ volume was 6.97 cc. Does overall size justify an image-based diagnosis of BPH?

Does BPH explain the overall enlargement of this patient’s prostate or his obstructive voiding symptoms?

The medical literature is replete with measures of total prostatic and TZ size. But the parameters by which we measure the PZ contribution to enlargement remain undefined. Prior authors have suggested subtracting the calculated TZ volume (TZV) from PTV to derive the volume of the PZ (PZV) [3-5]. TZ and PZ volumes have been used to evaluate the relationships between these volumes and urologic measures of bladder outlet obstruction e.g. uroflowmetry and symptom scoring [3,4,6,7]. However, there are few attempts to *directly* measure the thickness of the PZ band surrounding the TZ from imaging and none using MRI in symptomatic or asymptomatic patients with “normal” TPV [8,9,6].

There are very uncommon patients in whom the peripheral zone PZ is markedly enlarged on MRI in the absence of TZ BPH despite that impression on DRE. If total TPV exceeds established “normal”, this enlargement is often attributed to BPH even when the TZ is not obviously enlarged. Radiologists and urologists should be aware of this possibility when the DRE is suggestive and imaging reports confirm increased TPV.

The main purposes of this study is to report the precision of a technique of measuring the PZ thickness on T2-weighted prostatic MRI and report normal parameters in patients with normal-sized prostates. We also wanted to establish the mean and second standard deviations (2SD) above and below the mean as criteria for abnormally narrow or expanded PZ thickness. This knowledge may contribute to an understanding about the significance of PZ thickness when patients are evaluated for clinical lower urinary tract symptoms (LUTS) and perceived enlargement on DRE. A secondary purpose is to explore TZ measurement in this small volume cohort along with various formulae relating the TZ volume, to TPV and PZ volume suggested in the urologic literature. In addition, an example of the application of this information to the occasional patient with disproportionate increase in PZ volume and thickness relative to central TZ hyperplasia is presented. We believe that this knowledge can be applied in the interpretation of imaging in patients with enlarged prostates to determine to what extent enlargement is due to BPH and to what extent it is due to abnormal PZ thickness.

## Material and Methods

### Patient Selection

This was a retrospective review of our institutional medical records and picture archival system. The protocol was approved by our institutional review board and ethics committee. The initial study population was referred to our tertiary academic medical center for MRI because of possible adenocarcinoma of the prostate based upon abnormal serum PSA measurement and/or abnormal DRE. Of this 1566 consecutive cohort, 132 separate subjects with normal-sized prostates were included in this study. Mean age was 58.2 years (15-82). Median serum PSA was 6.2ng/mL (range, 0.3-145). Most were asymptomatic for lower urinary tract symptoms (LUTS). Median American Urologic Association symptom Score (AUASS) was 2.2 (minimum 0, maximum 26) on a scale of 0-35. Inclusion criteria in this study required technically adequate T2-weighted MRI and TPV ≤ 25 cc. Exclusion criteria included post prostatic surgical patients, those previously treated with radiation for cancer, patients having had medical management or minimally invasive therapy for BPH, those being treated for prostatitis, and patients with TPV greater than 25 cc as measured on MRI. Patients with suspected tumor expanding or obscuring measurement boundaries were also not considered.

### MRI Technique

Prostate MRI was performed at 3T with a pelvic phased array coil without (94) and with (38) endorectal coil using (Magnetom Skyra or TrioTim; Siemens Healthcare; Erlangen Germany) scanners. T2W FSE imaging was performed in the axial plane (TR/TE 3700/80 msec; NEX 3; 3mm slice thickness; no interslice gap; flip angle 160 deg; FOV 140 mm, matrix 320 ⨯ 256) and coronal plane (TR/TE 4030/100 msec; NEX 2; 3 mm slice thickness; no interslice gap; flip angle 122deg; FOV 180 mm; matrix 320 ⨯ 256). T2W 3D SPACE images were obtained (TR/TE 1400/101 msec; flip angle 135 deg; 256 ⨯ 256 ⨯ 205 matrix, 180 mm FOV). In addition to 1 mm axial images, T2W 3D SPACE images were reconstructed at 3 mm in the axial, sagittal, and coronal planes. Additionally, dynamic contrast enhanced T1-weighted (3 mm slice thickness; TR/TE 4.9/1.8 msec; 224 ⨯ 156 matrix, 250 mm FOV, temporal resolution <10 sec), and diffusion-weighted images at b values of 50, 800, and 2000 s/mm3 were performed.

### Measuring Techniques and Analysis

All images were reviewed on high resolution monitor PACS using Philips Intellispace v4.4 software. Total prostate volumes were measured using the biproximate prolate ellipsoid model (length x width x anteroposterior diameter x 0.52). Length measurement in the mid-sagittal plane made by drawing a perpendicular line from the vesico-prostatic junction (VPL) to the apex. Any prostatic tissue above the VPL was measured separately as a perpendicular from the proximal margin of the tissue to the VPL and added to the measurement below the VPL to derive total length of the prostate (Fig. 2). This procedure has been previously explained and validated for inter-rater reliability and compared with the traditional ellipsoid method of measuring length directly from the proximal prostatic margin to the apex [10]. TZ volumes were measured using the traditional landmarks for calculating volume using the ellipsoid formula. The mean of these volume measures from the two examiners were averaged to obtain the “pooled” total volumes of the prostate and TZ. Peripheral zone measurements were independently made from T2 MRI images by two board-certified diagnostic radiologists, one with over 40 years of prostate volume measurement experience and one early career with less experience using prostate volume metrics.

**Fig. 2.**
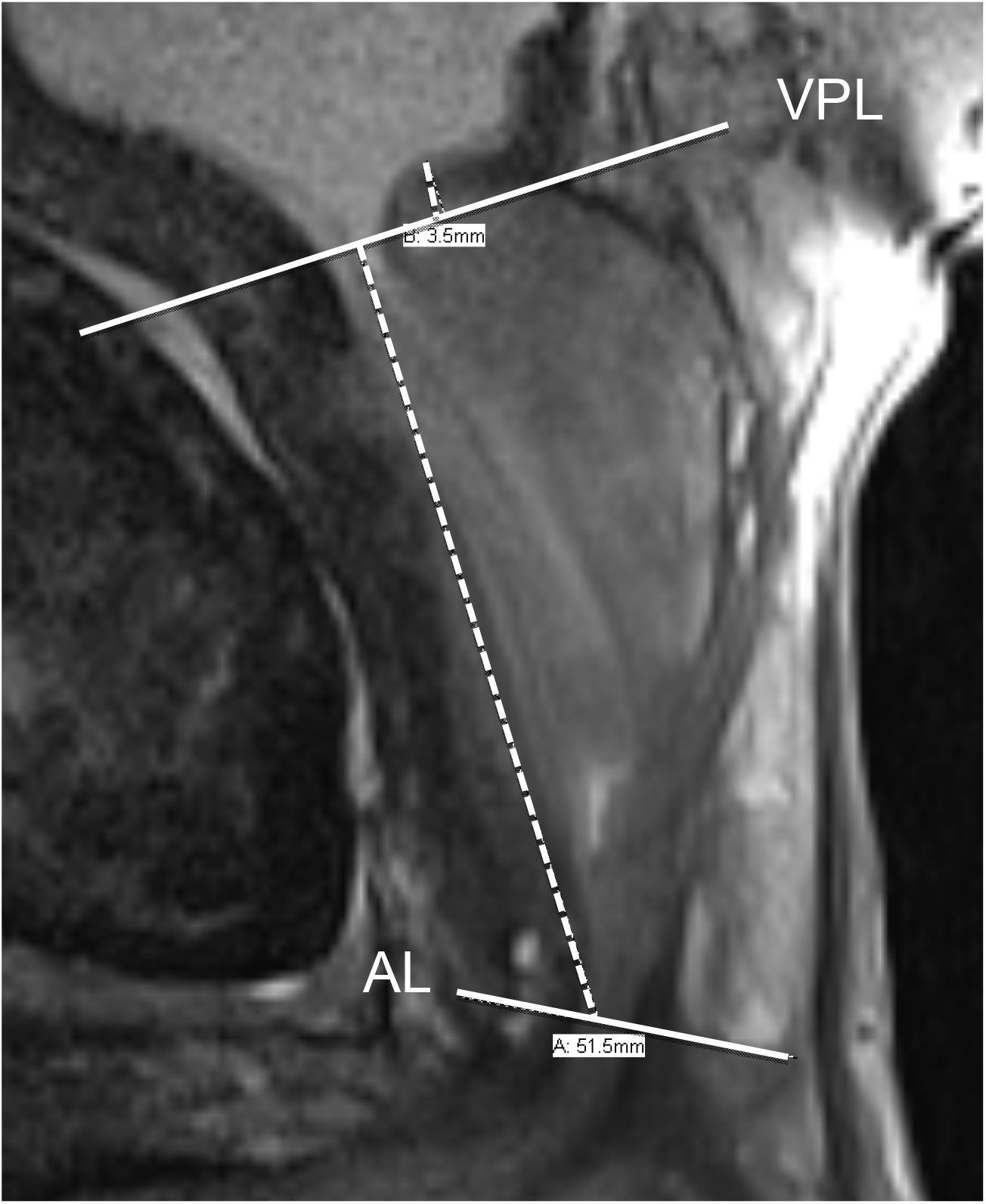
T2-weighted mid-sagittal MRI showing method of prostatic length measurement from apex to base. Measurements are made from apical line (AL) perpendicular to the vesico-prostatic line (VPL). If the prostate projects above the VPL, a line is made from the most proximal margin of the prostate perpendicular to the VPL and the two measures are summed to achieve the total length (55 mm).

The distribution of the PZ extends from the prostatic base to its apex. In these men with minimal TZ and/or retro-urethral BPH, PZ thickness measurements were made in the axial plane at the level of largest transverse diameter. Right and left posterolateral PZ (PLPZ) measurements were made from the estimated outer margin of the periurethral hyperplastic tissue to the inner boundary of the EPC following a perpendicular line defining the longest distance to the EPC (Fig. 3). The mean of the right and left sides in millimeters (mm) was determined for each examiner, and the average of the two readers’ means recorded as the “composite” (pooled) PLPZ thickness.

**Fig. 3.**
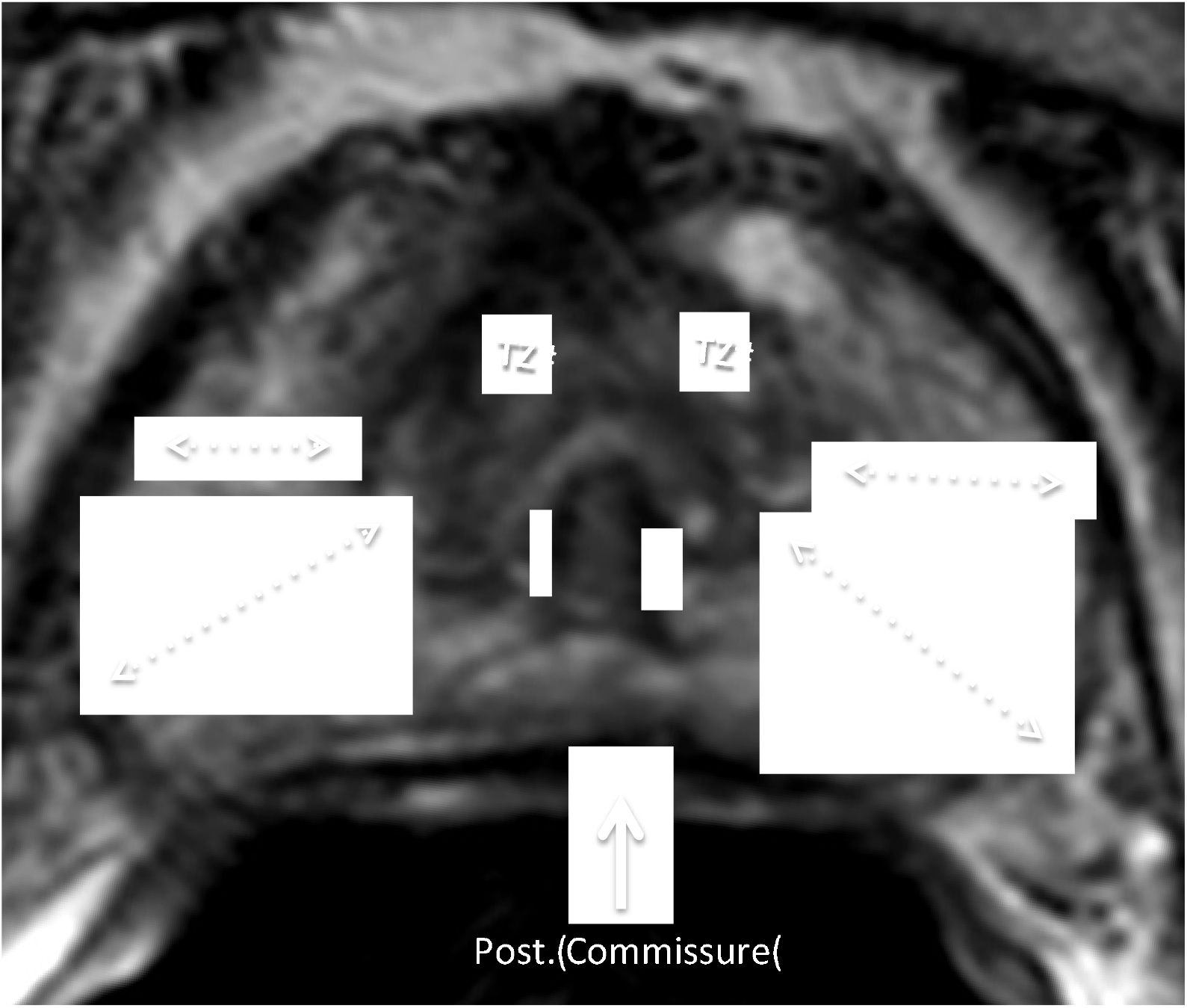
Axial plane of maximum transverse diameter showing lateral and postero-lateral peripheral zone (PLPZ) measurement boundaries.

In addition to measured variables, we analyzed calculated ones, particularly the transition zone index (TZI) representing the ratio of the TZ volume to TPV. PZ volume was calculated by subtracting TZ volume from TPV [3]. Other calculations included the ratios of the PZ volume to TPV, TZ volume to PZ volume, and PZ to TZ volume.

### American Urological Society Symptom Scoring

The medical records were reviewed for documentation of each patient’s symptoms. The American Urological Association Symptom Index (AUASS), a validated standard symptom-scoring instrument based on a seven questions regarding symptoms of the lower urinary tract [11] was found in 68 subjects. In the absence of AUASS documentation in the medical record, “no reported voiding symptoms” or “no symptoms” was taken to mean AUASS equivalent of zero score. For 59 patients who reported minimal symptoms in the record without formal scoring, we attempted to assign an appropriate score. Five patients had complete absence of AUASS or text mention of symptoms. The latter were not included in statistical evaluation of symptoms leaving 122 for statistics involving symptoms.

### Statistical Analysis

Statistical calculations were made using the QI Macro Statistics^®^ (KnowWare International, Inc., Denver CO, USA). Pooled individual observer data were used to calculate normal mean and 95% confidence intervals (CI) and 2SD limits on all PLPZ measurements for the entire final study group (Fig.4). Inter-rater concordance was calculated using the Pearson Product number (r). Conversions from r to two-tailed p-values utilized the vasserstats.net on line calculator. Since the distribution of the data was normal in most cases, the student t-test was applied using a standard of significance of <0.05. Similar calculations were obtained for stratified age groups and AUASS. Normal values and 95% CI were also documented for those with and without ERC. All variables were analyzed by ANOVA statistics at the 0.05 confidence level. The Kruskal-Wallis test was applied to one data set that was not normally distributed.

**Fig. 4.**
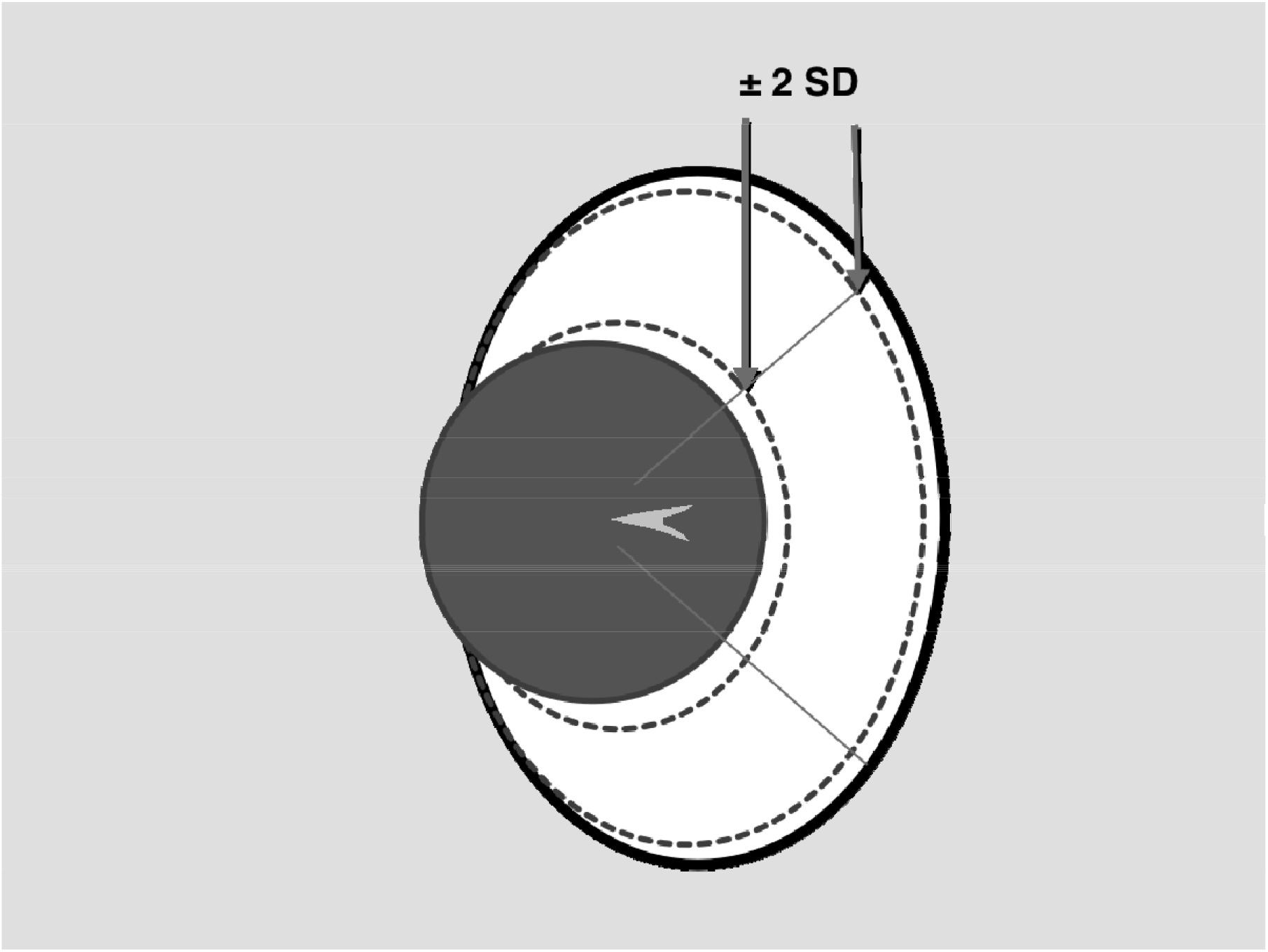
Solid straight lines are PLPZ measurement. Arrows point to dotted lines showing 95% upper and lower limits of normal (±2SD from mean) for right and left posterolateral peripheral zones (PLPZ).

## Results

### Peripheral Zone Thickness

Mean pooled (composite) measurement for the PLPZ was 10 mm (CI= 9.5-10.5 mm) with SD of 2.87 mm. Mean for the senior observer was 10.1 mm (CI=9.59-10.6) and for the junior observer 10.0 mm (CI=9.5-10.5). Pearson correlation between the two observers was 0.63 (p=0.001) with r^2^ =0.392. Two-sided Student t-test of observer mean measurement of PLPZ resulted in p=0.75. (Could not reject the null hypothesis, thus, the means were the same). An upper limit of normal PLPZ defined by the second standard deviation above the mean was 15.8 mm (Table 1).

**Table 1.** Measurement of PLPZ for 2 Observers

Measurement of PLPZ for 2 Observers
|  | Mean<br>(mm) | Min.<br>(mm) | Max.<br>(mm) | 95% CI | 2SD | Pearson<br>(r) | t-test<br>(p) |
| --- | --- | --- | --- | --- | --- | --- | --- |
| Senior | 10.1 | 1.8 | 22 | 9.6-10.6 | 4.2-16 |  |  |
| Junior | 10.0 | 2.3 | 20.8 | 9.5-10.5 | 4.4-15.6 |  |  |
| Pooled | 10.0 | 2.5 | 21.1 | 9.5-10.5 | 4.3-15.8 | 0.63<sup>†</sup> | 0.667* |

±2SD (in millimeters)
<sup>†</sup>=p<0.00001 \*Cannot reject the null hypothesis since p>0.05 (means are the same)

### Effect of Age on PLPZ

Mean PLPZ thickness by age is shown in table 2. ANOVA resulted in p=0.46 between groups indicating no significant difference among the means.

**Table 2.** PZ Thickness by Age

PZ Thickness by Age
|  | ≤49 yrs. | 50-59 yrs. | 60-69 yrs. | ≥70 yrs. | Total No. | ANOVA |
| --- | --- | --- | --- | --- | --- | --- |
| n | 28 | 39 | 41 | 24 | 132 |  |
| Mean | 10.3 | 10.1 | 10.0 | 9.3 | 10.0 | 0.462* |

\*Cannot reject the null hypothesis since p>0.05 (means are the same)

### Effect of ERC Use On PLPZ

Mean PLPZ without ERC was 10.5 mm and without ERC equaled 9.8 mm. 2-sided t-test resulted in p=0.30 indicating that the means were the same and that posterior external compression due to use of the ERC did not significantly reduced the mean PLPZ thickness.

### Total Prostatic Volume

Mean pooled measurement of prostate total volume was 20.5cc (CI=19.9-21.1). Mean for the senior observer was19.6 cc and for junior observer 21.4 cc. Kruskal-Wallis test for non-normal distributions resulted in, rejection of the null hypothesis at the < 0.05 level, indicating the means were different. However, Pearson correlation of total prostate volume was r=0.57 (p<0.01).

### Transition Zone Volume and Transition Zone Index

Mean pooled measurement of TZ volume was 6.6 cc (CI=6.2-7.0). Mean for the senior observer was 6.9 cc compared with a mean for the junior observer of 6.2 cc. Upper limits for normal (+2SD from mean) was 9.7 mm. Student t-test resulted in p=0.03 rejecting the null hypothesis (means are different). Pearson correlation of TZ volume was r= 0.44 (p<0.01). Results of measurements for TZI are shown in table 3. Pooled TZI was 0.32 resulting in Pearson r=0.27 (p<0.01).

**Table 3.** TZI (TZ index=TZ/Total prostatic vol)*

TZI (TZ index=TZ/Total prostatic vol)\*
|  | Mean | Min. | Max. | 95% CI | Pearson (r) | t-test (p) |
| --- | --- | --- | --- | --- | --- | --- |
| Senior | 0.36 | 0.01 | 0.84 | 0.33-0.39 |  |  |
| Junior | 0.28 | 0.00 | 0.72 | 0.27-0.30 |  |  |
| Pooled | 0.32 | 0.06 | 0.61 | 0.30-0.34 | 0.27 | <0.05 |

\*Means may be different. Dependent on accuracy of TZ and Total prostatic vol. measurement

### Other Measurements

PZ Volume showed r =0.37 (p<0.01), but the means were not the same between the two observers (table 4). Recorded AUASS scores numbered 66. There were 55 medical record-assigned scores based on text entries–all less than 7. No symptom scores could be found or estimated on 11 subjects. Median AUASS was 2.2 (range, 0–23). When grouped by age, there was no significant difference in mean AUASS (table 5). There was no meaningful correlation between AUASS and PLPZ, TPV, PZ volume, TZ volume, TZI, TZ/PZ ratio or PZ/TZ ratio.

**Table 4.**
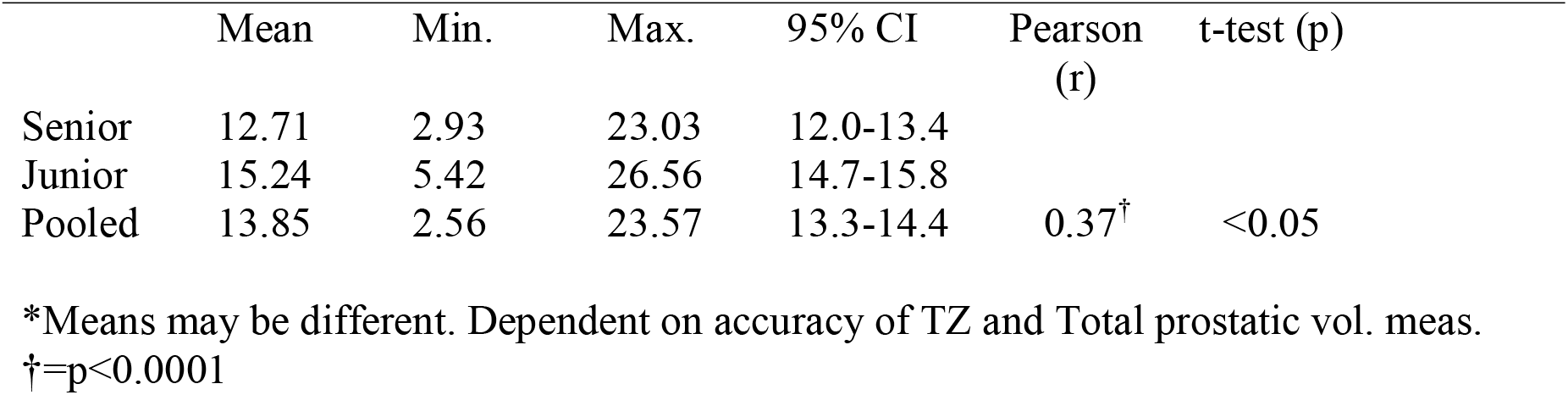
PZ Volume*

**Table 5.**
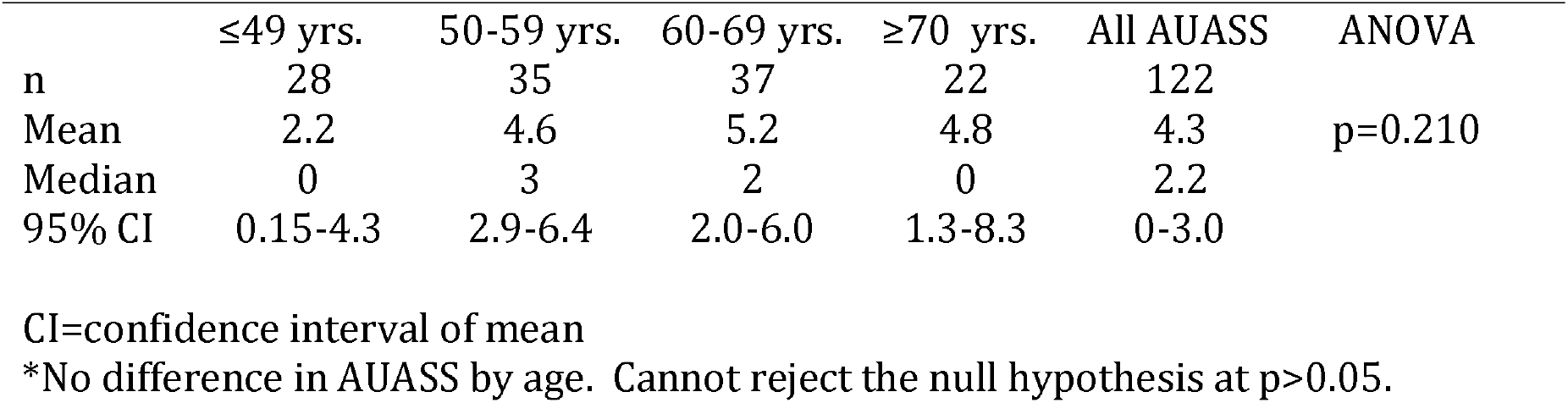
AUASS by Age*

## Discussion

Our main finding was determining the mean and 95% SD for PLPZ in normal-sized prostates according to our definition. We intended to learn if this measurement was significantly dependent on examiner experience. Pearson correlation coefficient of 0.63 (P<0.01) indicates no significant difference between the senior and junior examiner when measuring PLPZ thickness (Table 1). In addition, student t-test demonstrated the means were the same for both examiners (p>0.667). 95% lower and upper limits of normal for PLPZ of 4.3 and 15.8 mm were determined on MRI for the first time.

Precision of PLPZ thickness is in accord with the findings of Kwon, et al [9] who using TRUS and somewhat similar measurement techniques achieved higher inter-observer correlation (r=0.896) than was found in our study (r=0.57). This may be attributable to three factors: our differences in the experience of our 2 observers, our selection of a more difficult landmark (outer TZ margin versus urethra), difference in case selection (normal-sized prostates versus a spectrum of volumes. There is room for optimism that our measurement technique may not only possess inter-rater reliability (precision) but seems likely to be transferable to the general community of radiologists looking at a spectrum of abnormal-sized prostates with more discrete TZ margins. PLPZ measurement with the use of the ERC was the same as without. Therefore, our means and standard deviations values can be applied to both situations.

Our study confirms greater inter-reader correlation (precision) in the estimating the size of the PZ measuring PLPZ verses calculation of PZ volume. The use of this direct and easier measurement of PZ thickness is favored over the indirect calculation of PZ volume derived from subtracting TZ volume from TPV. This also obviates the difficulties of measuring the TZ length on lower resolution reconstructed MRI images in the calculation of TZV.

The first attempt we know of to measure thickness of tissues surrounding the prostatic urethra with was done by Suzuki T, et al using axial sections on 47 postmortem patients (8). Their technique was different from ours in two additional fundamental ways. They measured at 5 axial levels and measured PLPZ from the “posterior wall” of the urethra to the outside of the prostatic “capsule”. In the 40 patients with total prostate weight of <30 gm, the mean posterolateral distance measured 13.5 mm including what we would now term TZ tissues.

There are only two previous recorded measurements of PZ thickness in the medical literature using imaging. Kwon, et al, [9] utilizing transrectal ultrasound (TRUS) in a patient population with larger mean TPV and TZV of 33.3 cc and 16 cc respectively and a larger TZI of 0.45, found mean PZ thickness to be 11.1 mm and 2 standard deviations (2SD) above the mean of 16.1. This is similar to the 15.8 in our cohort of normal-sized prostates. Their report compared clinical symptoms and urodynamic studies in 3 cohort groups defined by PLPZ thickness measurements of <9.5 mm, ≥9.5-<13mm, and >13 mm. TRUS-measured PLPZ thickness correlated with symptom scores and all uroflowmetry measurements. They showed that as PLPZ narrows, symptom scores rise and there is an association with worsening uroflowmetry findings. PZ thickness showed independent correlation with symptom score, quality of life score and uroflow studies [9]. The latter urged further studies to see if PZ thickness can act as a proxy for pressure-flow studies in the evaluation and management of clinical BPH. Our results using AUASS as a marker for obstruction in normal sized prostates do not confirm those of Kwon in patients with enlargement. Matsugasumi, et al [6], using MRI segmentation measured PZ thickness (PZT) at a *median* value of 12.8 (range 2.2-24.6 mm) but provided no mean or standard deviation information. They concluded that there was negative correlation of PZT with symptom scores, TPV, TZV, or PCAR (ratio of maximal anteroposterior to transverse diameter). Matsugasumi and others have concluded that the thickness of the PZ does not change with age [6,5]. However, others have shown that PZ volume increases, but more slowly than TZV [4].

TZI has been actively discussed in the urologic literature. Kaplan, et al [12], using TRUS, was first to suggest a strong correlation between TZI to patient symptoms and significant urodynamic abnormalities. TZI >0.50 has been reported to be associated with symptoms and signs of BPH [12]. Some suggest a cut point of >0.40 as predictive of clinical BPH [13]. Others have not been able to demonstrate these associations [14, 15, 3]. The mean pooled TZI of 0.31 in our study is consistent with our population’s normal prostate volume and low median total AUASS of 2.2. One limitation of our study is inaccuracy by allowing for author assigned estimated symptom scores in patients without clear medical record documentation. Another limitation was the measurement of TZ volume. Wities, et al. [7] alluded to this as a problem in a population averaging over 40 gm in TPV and mean TZV of 22 gm. Patients with normal-size prostates and small TZ volume show indiscrete margins of the TZ since growth has not yet caused compression and thickening of their outer boundary (sometimes referred to as the “surgical capsule”). Identifying and measuring those boundaries in three planes can be very challenging, especially in planes that are reconstructed images. “Fuzzy” measurement will inevitably effect the computations of the other ratios that use TZ volume to characterize the prostate. Our data confirms this limitation. Despite this difficulty, our inter-observer correlation was 0.44 (p<0.01). However, the student t-test showed the means to be significantly different.

There was a significant difference in mean TPV between the two observers. This may be explained by the fact that while the senior observer was very experienced with the technique, the junior observer was new to the use of the alternative length measurement. Another explanation for the disparity could be that the junior reader was insufficiently tutored in the application of the TPV measuring method since emphasis of this report was on measuring the PZ.

Notwithstanding TZV measurement difficulties, mean pooled TZ volume was 6.6 cc in our study of men with TPV ≤ 25. This compares with mean volume of 10.6 cc in a Japanese screening study of asymptomatic men over age 55 years [16] and 9-15 cc in a population screening of Chinese men 40-70 years of age [17]. However, TZV reported in most studies in the United States and Asia in men of average age over 55 years ranges from 21-26 cc in subjects with TPV averaging 35-49 cc [6, 7, 15, 18,19,11,20,21]. Since TZV is known to correlate to TPV our results come as no surprise [22,4].

In addition to the TZI, it must be remembered that TZ volume is subtracted from TPV to obtain the volume of the PZ. PZV for prostates in our study ≤ 25 cc was 13.9 cc. compared to much higher values (usually 35-50 cc) in most other studies where patients are selected under different criteria e.g. BPH, LUTS, or enlargement on DRE.

Pooled PZ Volume of 13.9 is in general accord with other studies of men with prostates of low volume. Fujikawa, et al [23] states that “Generally, the prostate grows and develops rapidly, mainly in the PZ, during adolescence, peaks in 20s and starts to shrink in the late 40s, which coincides with frequent adenoma development in the TZ.” A similar trajectory for PZ thickness showing relative stability after middle age was demonstrated with MRI [5,6].

Pooled TZ/PZ volume ratio of 0.58 can be compared with the findings of Meikle, et al [4]. Pooled TZ/PZ volume of 0.58 showed Pearson r=0.20 (p<0.02). This is an interesting and unexpected finding. Meikle [4] found that patients with AUA symptom scores <10 showed a mean ratio of TZ volume to PZ volume of 0.37. Patients with mean total volume 31.2 ±2.6 cc and symptom scores ≥10 had TZ/PZ ratio above 0.67. (Derived from Meikle Table 2) [4]. Our largely asymptomatic patients with mean pooled total volume of 20 cc had TZ/PZ ratios closer to those who would have been expected to be more symptomatic according to the latters’ data.

PZ/TZ ratio in our study was 2.56 showing r=0.17 (p=0.051) with the same means for the two examiners. This is in accord for less symptomatic patients in which the PZ/TZ ratio was 2.7 in the study by Miekle, et al. [4].

Symptom scores demonstrated no correlation to PLPZ, TPV, TZV, PZV, TZI or other TZ to PZ ratios. There was also no correlation to AUASS by age category (Table 5). Because of the significant use of author assigned symptom scores, no definite conclusions should be made from this data.

Limitations of our study begin with its retrospective and cross-sectional nature. Generalizations about zonal growth, including the PZ have never been subjected to longitudinal prospective growth measurements in individual subjects. All assumptions in past or present studies are based on cross-sectional research. Measurement of the TZ is difficult, even with MRI, calling into question measurements and calculations involving the TZ in this study including PZV and TZI. Greater measurement training for the junior reader in this experiment could have improved the inter-rater reliability of all measurements, but the authors were most interested in how well the PZ thickness measurement technique might be expected to perform in community practice by radiologists without intensive preliminary training. Selection of PZT measurement was arbitrarily done at the axial level of greatest transverse size because of likelihood of greatest reproducibility. It is possible that this is not the optimal axial level for such measurement. While few of the patient cohort had biopsy confirmed cancer, it is possible that tumor may have had minor influence on some measurement boundaries.

The PLPZ for the opening introductory case showed measurements outside the 4^th^ SD form the mean (Fig. 5). Therefore, the overall enlargement of the prostate is not due to mechanical effects of BPH, since TZ volume is minimal. Obstructive symptoms could be attributed to dynamic causes. The PZ enlargement is likely incidental. The exact etiology and pathogenesis for disproportionate PZ enlargement is unknown but is being studied.

**Fig.5.**
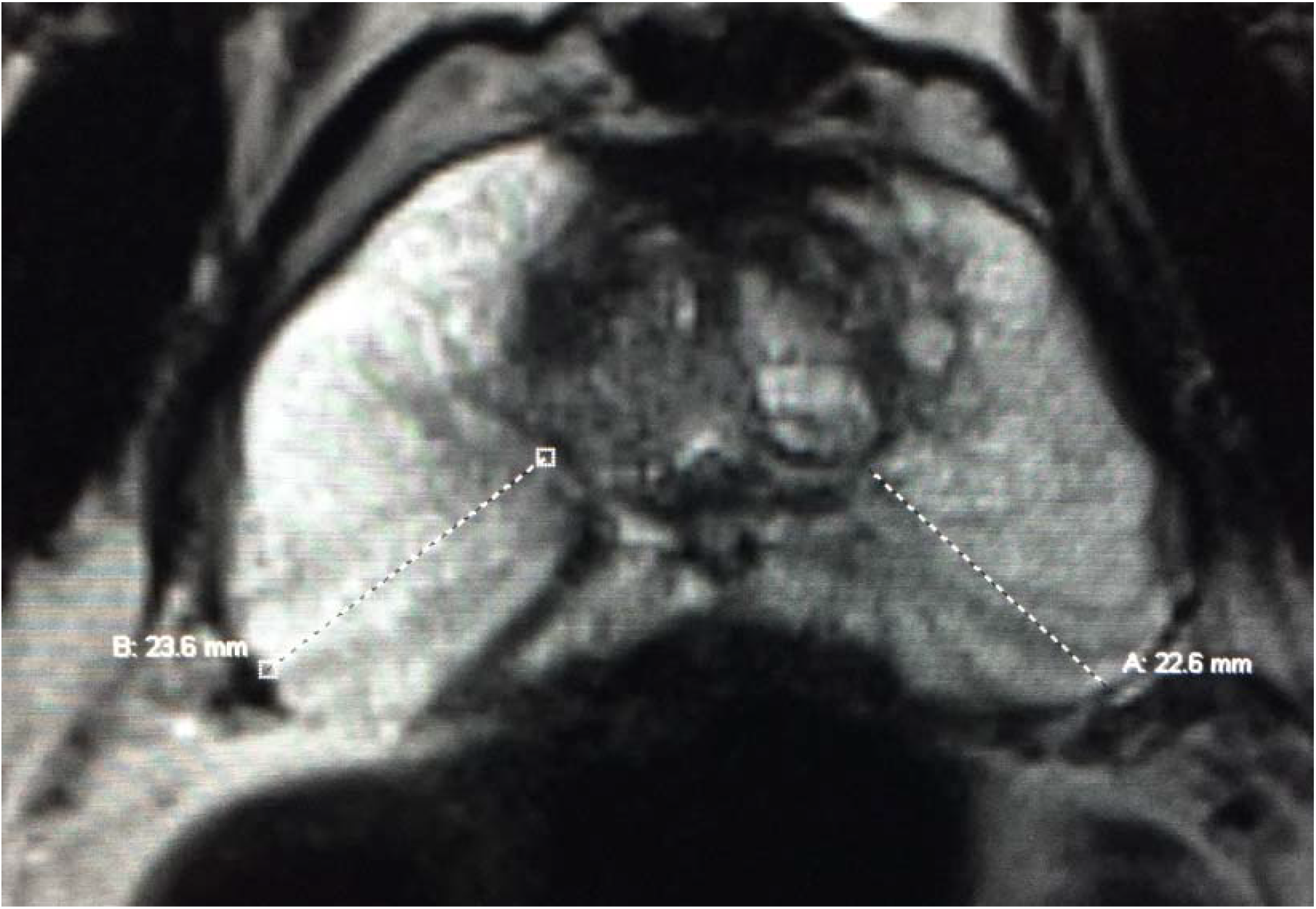
T2-weighted MRI measuring PLPZ shows thickness outside the 4^th^ SD from the mean and relatively little TZ hyperplasia. This represents PZ enlargement without BPH. Though the total volume of the prostate exceeds 35 cc, this should not be diagnosed as BPH on imaging. Biopsies negative for carcinoma.

## Conclusions

1. Precision of the PZ thickness measurement technique was demonstrated.
2. Normal values for mean posterolateral PZ thickness including 95% upper and lower thresholds were measured in “normal-sized” prostate glands ≤ 25 cc in a cross-sectional retrospective study. Mean pooled (composite) measurement for the normal PLPZ is 10 mm (CI= 9.5-10.5).
3. PZ thickness in excess of 15.8 mm defines abnormal PZ enlargement, while PZ thickness less than 4.8 mm defines compression.
4. Like PZV, PLPZ thickness is independent of age and use of ERC. Because the PZ changes little in size after the late 40s, these normal values should be useful to apply in patients of all ages.
5. Direct measurement of PZ thickness seems easier and more reproducible (r=0.63) than indirect calculation of PZV (r=0.37) in normal size glands. Whether this is true in glands with larger BPH remains speculative.

## Data Availability

The data for this study is available for any reasonable request.

## Notes

The authors have no conflicts of interest to report.

### Competing Interest Statement

The authors have declared no competing interest.

### Clinical Trial

NA

### Clinical Protocols

https://www.abdominalradiology.com

### Funding Statement

This study was not funded.

